# How does spinal movement variability change in people with low back pain?: protocol for a systematic review

**DOI:** 10.1101/2020.04.07.20053538

**Authors:** Hiroki Saito, Hiroshi takasaki, Yoshiteru Watanabe, Toshiki Kutsuna, Toshihiro Futohashi, Masayoshi Kubo, Yasuaki Kusumoto, Hiroki Tiba

## Abstract

Low back pain(LBP) is the number one cause of disability worldwide. One factor which might potentially contribute to ongoing pain is impaired spinal movement variability. It is uncertain how movement variability changes during trunk movements in the presence of LBP. In this protocol, we will systematically investigate and compare both the amount and structure of spinal movement variability during repeated trunk motions between people with and without LBP. The results will be reported in line with the PRISMA(Preferred Reporting Items for Systematic Review and Meta-Analysis). Searches will be conducted on CENTRAL, PubMed, MEDLINE, EMBASE, and CINAHL databases, along with a comprehensive review of grey literature and key journals.Three pairs of two independent reviewers will screen potential studies and two independent reviewers assess the risk of bias within studies which meet the inclusion criteria. The Newcastle-Ottawa risk of bias tool will be used to assess the quality of the data.

## Background

Low back pain(LBP) is the number one cause of disability worldwide.^1^ Although there are multifactorial contributors to disable LBP including psychological, genetic and social factors, biophysical factors are demonstrable.^2^ One of the fundamental bases of biophysical factors is that there is an association between LBP and aberrant motor control strategies ^3–5^. This includes impaired activity patterns of abdominal and back muscles ^6^, altered range of motion^7^ and proprioception deficits. ^8^

There has been an increasing interest in human movement variability that is defined as the natural variations in motor control strategies.^9^ Human movement system obtains multiple degrees of freedom using variable muscle recruitment patterns, joints and nervous systems that allow the different solution to a particular task ^10,11^. Thus, variability is a necessary condition for maintaining or achieving functional movement, providing flexible and adaptive strategies that do not depend on rigid programs for each task.^12^ Furthermore, optimal movement variability can prevent musculoskeletal injuries by sharing the load across the human body. ^9,13–15^

It is thought that impaired movement variability is associated with the onset or persistence of pain.^9,15^ One considers decreased variability may reflect the constraint of movements, causing repetitive stress on tissues. ^13,16^ Other considers increased variability may reflect an inconsistent or unstable coordination pattern. ^17,18^ This means that mid-range alignment of segments could be compromised, which may lead to higher tissue strains. ^19–21^ Thus, there could be some degree where healthy individuals move functionally and out of this window (i,e. too high variability or too low variability) individuals may develop injuries.^15^

It is uncertain how movement variability changes during trunk movements in the presence of LBP. Investigating altered spinal movement variability in people with LBP will uncover new directions for understanding motor control deficits that may cause or persist LBP. There are two aspects that assess movement variability. The one concept, which represent the amount of variability, is to quantify using linear statistical tools (e.g. range, standard deviation, and coefficient variation) while another concept, which represent the structure of variability, is to quantify using non-linear tools (e.g. percent recurrence, percent determinism, sample entropy and Lyapunov exponent).^12,22,23^ They are not functionally dependent of one another.^12^ We will, therefore, design the review to systematically investigate and compare both the amount and structure of spinal movement variability during repeated trunk motions between people with and without LBP.

## Methods

### Protocol, search strategy, and study selection

We will search the following databases(from inception to 16 January 2020): CENTRAL, Medline, Pubmed, Embase, and CINAHL. There was no date or language restrictions. The following inclusion and exclusion criteria will be applied to the articles.

Inclusion criteria

- The study must have been published or ‘in press’ prior to 16 January 2020
- Studies conducted on humans
- Studies conducted on adults
- Studies that included people with non-specific LBP and control(without LBP)
- The study must measure the spinal angle relative to pelvis or thigh during a repeated trunk movement and assess either the amount of variability or the structure variability
- The study must include both the LBP group and the control group.

### Exclusion criteria

- Duplicated publications
- Studies that included people with serious pathology(eg, fracture, infection, cancer, central nerve system disease, respiration disorders) or pregnant and having given birth within 3 months.
- Studies without the control group
- Cadaveric or animal studies
- Single-subject case reports
- Unpublished articles

In addition to electronic search strategies,searching potentially relevant studies from other sources was performed:

1. Reference searches of relevant reviews, including previously published systematic review of kinematic changes and LBP ^7,24–27^
2. Review of personal files of investigators, which included authors of previous focused reviews of movement variability ^9,12,15^

Three pairs of assessors(HS & YK, HT & TF, YW & TK) independently will screen the titles/abstracts of all retrieved records for apparent exclusions and then apply our inclusion criteria to the full text of the remaining papers. Disagreements will be resolved by discussion between these examiners in order to achieve consensus on which studies to be included.

### Data extraction

Data extraction will be carried out by the two independent reviewers (HS, YK). Predetermined data extraction included general study information (author, year of publication); study design and study population details(LBP, control); year and methods of data collection; and outcomes.

After data extraction, the final inclusion and exclusion decisions will be made by the consensus from the two reviewers(HS, YK) regarding the study, according to the predetermined criteria. The information is detailed in table format to compare and contrast the study designs, objective, LBP/control population measurement, and outcome measure.

### Risk of bias in individual studies

Two independent reviewers(HS, YW) will appraise the included studies using the Newcastle-Ottawa Scale (NOS).^28^ The disagreement will be resolved with discussion. The NOS is useful for both case-control and longitudinal (prospective studies) and evaluated three quality parameters (selection, comparability, and outcome) divided by eight items. ^28,29^ The items of selection and outcome are scored from one point while the item compatibility can be scored up to two points adopted to the specific topic of interest. ^28,29^ Similar to the previous review, ^24^ two items of the method for data acquisition(i.e validity of measurement tools and reliable data acquisition) will be added. Thus, 8 items were assessed using NOS in which the maximum for each study was 9 and other 2 items regarding the data acquisition will be also assessed separately.

## Data Availability

We intend to publish the review on completion

